# Neighborhood distribution of availability of newer tobacco products: A four-site study, 2021

**DOI:** 10.1101/2022.05.07.22274794

**Authors:** Shyanika W. Rose, Arati Annabathula, Susan Westneat, Judy van de Venne, Mary Hrywna, Christopher Ackerman, Joseph G. L. Lee, Mahdi Sesay, Daniel P. Giovenco, Torra Spillane, Shawna V. Hudson, Cristine D. Delnevo

## Abstract

**Background:** Audits of tobacco retail stores can identify marketing patterns as newer tobacco products are introduced in the US. Our study examined store and neighborhood correlates of availability of nicotine pouches and disposable e-cigarettes in four US sites.

**Methods:** We conducted standardized store audits of n=242 tobacco retailers in 2021 in different states: New Jersey, Kentucky, North Carolina, and New York. Store audits focused on availability of nicotine pouches and disposable e-cigarettes. We geocoded stores linking them with census tract demographics. We conducted unadjusted and adjusted Poisson regression of availability of each product with correlates of the proportion of Non-Hispanic White residents, households under poverty, proximity to schools, site, and store type.

**Results:** Nicotine pouches and disposable e-cigarettes were each available in around half the stores, but availability differed across sites. In adjusted analyses, nicotine pouches were less likely to be available in each store type vs. chain convenience and more likely in stores in census tracts with more non-Hispanic White residents. In contrast, disposable e-cigarettes were more likely to be available in tobacco store/vape shops than convenience stores and less likely in non-specialty store types like groceries.

**Conclusions:** The availability of newer tobacco products like nicotine pouches and disposable e-cigarettes were widely available in stores across sites, but retail marketing patterns appear to differ. As these product types become subject to increased regulation as they go through the FDA pre-market authorization process, understanding changes in their retail environment is critical to inform potential policies regulating their sale and marketing.

## Introduction

Retail outlets are one of the main avenues for marketing and promotion of tobacco products in the US. In 2020, cigarette manufacturers spent $7.84 billion to advertise and promote tobacco – almost 80% for discounts and other promotions at the point-of-sale.^1^ Exposure to point-of-sale marketing of tobacco products has been linked with youth and adult tobacco use and unsuccessful quit attempts among smokers.^2,3^ There is also clear evidence of tobacco manufacturer targeting of products and marketing by neighborhood demographics with substantial consequences for health equity.^4^ A higher density of tobacco retailers have been found in neighborhoods with higher proportions of Black and low-income residents, potentially contributing to disparities in tobacco product use.^5^ Additionally, different products have been disproportionately marketed in different communities. For instance, more retail menthol cigarette and cigarillo marketing is found in neighborhoods with more Black residents;^6,7^ however, smokeless tobacco marketing is generally found to be lower in neighborhoods with more non-White residents.^8^ Prior research on retail availability has shown that e-cigarettes were not initially marketed in neighborhoods with more residents from racial/ethnic minority backgrounds but became more prevalent in such neighborhoods over time.^9^

Retail surveillance of the tobacco product landscape can help identify early trends in newer tobacco product availability and marketing in communities. Several non-combustible product types including disposable e-cigarettes and smokeless nicotine pouch products have emerged on the US market, but little is known about retail availability of these products. In 2021, disposable e-cigarettes such as Puffbar were used by 54% of youth e-cigarette users after these products were exempted from flavor restrictions imposed on cartridge/pod-based e-cigarettes such as JUUL.^10^ Nicotine pouches, often marketed as ‘tobacco-free’ or ‘tobacco-leaf free’ nicotine, are newer smokeless tobacco products that come in pouches like moist snuff or snus but contain a nicotine powder concentration instead of tobacco leaf. Market leaders like Zyn and On! are sold by major tobacco manufacturers.^11^ Sales of such products have greatly increased since their introduction to the market in 2016.^11^

Understanding the retail availability of these products can help to determine to whom and where these products are marketed. However, to date, little is known about the retail availability and neighborhood and store correlates of recently introduced products. Thus, we sought to assess neighborhood differences in the availability of nicotine pouches and disposable e-cigarette devices as part of a four-state study investigating tobacco product availability and health inequity.

## Methods

### Data Collection

#### Data Source

Each site randomly sampled stores from tobacco retailer lists. Sites in New Jersey (NJ) and New York (NY) used tobacco retailer license lists. In Kentucky (KY) and North Carolina (NC), which lack tobacco retailer licensing, this came from compliance check lists used by the state for Synar compliance in KY and from a list of probable tobacco retailers created using a validated method in NC.^12^ In NC and KY, vape shops were identified using a validated search of Google Maps and Yelp.^13^ In NJ, vape shops were also added through Google search as these store types were not included on tobacco retailer license lists.

#### Sample

NJ included 65 stores selling tobacco, which were visited in prior rounds of data collection from stores in a 25-mile radius around Rutgers University campus center in New Brunswick, NJ and 10 additional vape stores. In sites selecting a new sample, each site randomly selected 50 tobacco retailers and up to 10 vape shops or as many as available in the jurisdiction. Thus, in these jurisdictions, stores were randomly selected from retailers in Fayette County, KY, (in the Lexington metro area) and Pitt County, NC, around Greenville, and the Borough of Manhattan in New York City, NY. Thus, the full sample set was comprised of KY (n=60), NC (n=56), NY (n=56), and NJ (n=74), bringing the total number of stores where audits were attempted to 246. During audits, we found 4 stores did not carry any tobacco and were thus ineligible and dropped from the analytic sample (n=242).

#### Store audits

Following standard store audit practices,^14^ trained data collectors at each site visited stores to record the different types of products sold and store characteristics. A Qualtrics survey was created with standardized questions used across sites. Each data collector used a smartphone to access this survey either while inside the store or while in the parking lot immediately after the store visit. The Qualtrics survey included a geolocator which identified the data collector’s latitude/longitude based on the phone location when completing the survey. Store auditors were trained to conduct audits covertly and not interact with store staff unless necessary. If the store did sell tobacco but was not safe or closed at the time of visit, the data collector returned to complete the audit at a later time. No products were purchased as part of the audit, but occasionally in smaller stores data collectors would make small purchases to make the visit appear more natural and avoid suspicion. These availability audits were conducted as part of a larger study dealing with underage purchasing of tobacco products. Audits were completed between April 2021 and September 2021.

#### Measures

We assessed availability (yes/no) of cigarettes, e-cigarettes (pod and disposable), cigars, smokeless tobacco (e.g., chew, dip, snuff), and tobacco-free nicotine pouches. Where data collectors responded yes to e-cigarettes, we followed up with “Which e-cigarette brands are available?” and gave visual examples and yes/no options for Puff Bar Disposable, Hyppe bar disposable, Posh Plus disposable, Fruyt disposable, Flair disposable, Pop disposable, Lava pod disposable, Eon smoke disposable, and a free-text response for other disposable e-cigarette brands. A yes to any of these items was coded that disposable e-cigarettes were available. Availability of JUUL or VUSE Alto tobacco or menthol pods was coded as pod e-cigarette availability. We coded store types into 10 categories (convenience store, drug store, gas kiosk only, dollar store, grocery/supermarket, mass merchandiser, chain convenience, vape shop, tobacco store, other store type).

#### Geocoding

After collecting product availability (see Table 1), ArcMap and ArcGIS Pro were utilized to geocode store locations based on address and cross-checked with the geolocation position collected in the field. If the points did not match, we would search coordinates on Google map to update the store address if needed. This verified geocoded position was used to identify the census tract of the store and merge stores with the most recent publicly available American Community Survey 5-year estimates (2015-2019) census tract data as well as the National Center for Educational Statistics’ public school (2020-2021) dataset. We performed a closest facility analysis to determine whether the store was within a half mile of a school. We picked this buffer to account for reasonable walking distance from schools in the areas that were less urban in our sample. We linked tract demographics (percent non-Hispanic White residents, percent households under federal poverty level) to stores based on their census tract.

**Table 1.** Product availability and store and neighborhood characteristics in tobacco retail audits in four sites, 2021 (n=242)

|  | KY<br>n=59 | NC<br>n=56 | NJ<br>n=71 | NY<br>n= 56 | Overall<br>n=242 | p-value |
| --- | --- | --- | --- | --- | --- | --- |
| **Product Availability** |  |  |  |  |  |  |
| Cigarettes | 49 (83%) | 51 (91%) | 62 (87%) | 49 (88%) | 211 (87%) | .4764 |
| Pod E-cigarettes | 33 (56%) | 19 (34%) | 30 (42%) | 37 (66%) | 119 (49%) | .0147 |
| Disposable e-cigarettes | 27 (46%) | 18 (32%) | 34 (48%) | 35 (63%) | 114 (47%) | .0150 |
| Cigars | 48 (81%) | 52 (93%) | 65 (92%) | 36 (64%) | 201 (83%) | .0004 |
| Smokeless tobacco | 43 (73%) | 22 (39%) | 28 (39%) | 19 (34%) | 112 (46%) | <.0001 |
| Tobacco-free nicotine pouches | 45 (76%) | 26 (46%) | 26 (37%) | 24 (43%) | 121 (50%) | <.0001 |
| **Store Characteristics** |  |  |  |  |  |  |
| **Store type** |  |  |  |  |  | <.0001 |
| Convenience Store (non-chain) | 4 (7%) | 14 (25%) | 30 (42%) | 34 (61%) | 82 (34%) |  |
| Chain Convenience | 30 (51%) | 11 (20%) | 16 (23%) | 2 (4%) | 59 (24%) |  |
| Drug store | 3 (5%) | 5 (9%) | 6 (8%) | N/A | 14 (6%) |  |
| Gas Kiosk only | 0 | 2 (4%) | 5 (7%) | 1 (2%) | 8 (3%) |  |
| Dollar store | 4 (7%) | 11 (20%) | 1 (1%) | 0 | 16 (7%) |  |
| Grocery store/supermarket | 6 (10%) | 5 (9%) | 1 (1%) | 0 | 12 (5%) |  |
| Mass Merchandiser | 2 (3%) | N/A | 0 | 0 | 2 (0.8%) |  |
| Vape Shop | 10 (17%) | 4 (7%) | 10 (14%) | 4 (7%) | 28 (12%) |  |
| Tobacco Store | 0 | 4 (7%) | 0 | 13 (23%) | 17 (7%) |  |
| Other store type | 0 | 0 | 2 (3%) | 2 (4%) | 4 (2%) |  |
| **Store Neighborhood Demographics** |  |  |  |  |  |  |
| **Non-Hispanic White residents** |  |  |  |  |  |  |
| Q1 (0-28%) | 2 (3%) | 15 (25%) | 27 (44%) | 17 (28%) | 61 (25%) | <.0001 |
| Q2 (28-60%) | 6 (10%) | 19 (32%) | 21 (35%) | 14 (23%) | 60 (25%) |  |
| Q3 (60-74%) | 22 (37%) | 21 (35%) | 8 (13%) | 9 (15%) | 60 (25%) |  |
| Q4 (74% +) | 29 (48%) | 1 (2%) | 15 (25%) | 16 (26%) | 61 (25%) |  |
| **Households under poverty** |  |  |  |  |  |  |
| Q1 (0-7%) | 17 (28%) | 0 | 35 (57%) | 9 (15%) | 61 (25%) | <.0001 |
| Q2 (7-17%) | 14 (23%) | 3 (5%) | 16 (27%) | 27 (45%) | 60 (25%) |  |
| Q3 (17-26%) | 15 (26%) | 22 (38%) | 10 (17%) | 11 (19%) | 58 (24%) |  |
| Q4 (26% +) | 13 (21%) | 31 (49%) | 10 (16%) | 9 (14%) | 63 (26%) |  |
| **Within 0.5 mile of school** |  |  |  |  |  | <.0001 |
| No | 36 (61%) | 42 (75%) | 22 (31%) | 0 | 64 (26%) |  |
| Yes | 23 (39%) | 14 (25%) | 49 (69%) | 56 (100%) | 178 (74%) |  |

#### Data Analysis

We conducted descriptive statistics by site using Pearson chi-square tests to examine differences in proportions across sites. Since the availability of nicotine pouches and disposable e-cigarettes was high in our sample, we conducted Poisson regression to estimate prevalence with robust standard errors to derive risk ratios.^15^ We then used unadjusted and adjusted Poisson regression analyses to examine the prevalence of (1) nicotine pouches and (2) disposable e-cigarettes separately. Initial results including the proportions of Hispanic and Black/African-American residents and median household income and percent households under poverty did not converge due to multicollinearity as these measures were correlated at α=0.76. Instead, for neighborhood measures we included percent of non-Hispanic White residents and percent households under poverty as well as a proximity measure of whether the retailer was within ½ mile of a school. Thus, adjusted analyses included these measures along with store type collapsed into four categories (chain convenience (ref), non-chain convenience, tobacco/vape stores, other store types [including grocery/supermarket, pharmacy, dollar stores, mass merchandisers, and gas kiosks]), and site (NJ (ref), KY, NC, NYC). We used Stata v16 and SAS 9.4 and conducted analyses in February 2022. This research was classified as Not Human Subjects Research by the Institutional Review Board at the University of Kentucky.

## Results

As shown in Table 1, almost all retailers carried cigarettes and cigars, but half or less of all retailers carried other products including pod e-cigarettes, disposable e-cigarettes, smokeless tobacco and nicotine pouches. There were significant differences in the availability of product types across sites except cigarettes. There were more stores in census tracts with the highest quartile of non-Hispanic White residents in Kentucky than in the other sites, while there were more stores in areas with the highest quartile of residents under poverty in North Carolina than other sites.

### Nicotine Pouches

As shown in Table 2, in unadjusted analyses, prevalence of nicotine pouches was significantly lower in all store types relative to chain convenience stores, in stores in census tracts with the 3^rd^ quartile of residents under poverty compared with the lowest quartile, and in stores near schools. Additionally, unadjusted prevalence of nicotine pouches was higher in Kentucky compared with New Jersey, and in neighborhoods with more non-Hispanic White residents compared with neighborhoods with the fewest non-Hispanic White residents. In adjusted analyses, only store type and the proportion of non-Hispanic White residents remained significant.

**Table 2.** Correlates of availability of Nicotine Pouches and Disposable E-cigarettes in tobacco retail stores in four sites (n=242)

|  | Product Availability |  |  |  |
| --- | --- | --- | --- | --- |
|  | Availability of<br>TFNP (unadjusted)<br>IRR (95% CI) | Availability of<br>TFNP (adjusted)<br>IRR (95% CI) | Availability of<br>disposable e-cig<br>(unadjusted)<br>IRR (95% CI) | Availability of<br>disposable e-cig<br>(adjusted)<br>IRR (95% CI) |
| **Store Type** |  |  |  |  |
| Chain Convenience store | ref | ref | ref | ref |
| Non-chain convenience store | **0.2 (0.2-0.4)** | **0.3 (0.2-0.4)** | 0.9 (0.7-1.3) | 1.0 (0.7-1.5) |
| Tobacco Store/Vape Store | **0.6 (0.5-0.8)** | **0.6 (0.4-0.8)** | **2.0 (1.5-2.6)** | **1.9 (1.5-2.6)** |
| Other Store types<br>(grocery/supermarket,<br>pharmacy, dollar store,<br>mass merchandiser, gas<br>kiosk) | **0.4 (0.3-0.6)** | **0.5 (0.4-0.7)** | **0.2 (0.1-0.4)** | **0.2 (0.1-0.5)** |
| **Site** |  |  |  |  |
| NJ | ref | ref | ref | ref |
| KY | **2.1 (1.5-2.9)** | 1.2 (0.9-1.7) | 1 (0.7-1.4) | 1.1 (0.7-1.4) |
| NC | 1.3 (0.8-1.9) | 1.3 (0.8-2.0) | 0.7 (0.4-1.1) | 0.9 (0.5-1.5) |
| NYC | 1.2 (0.8-1.8) | 1.5 (1.0-2.4) | **1.3 (1.0-1.8)** | 1.1 (0.8-1.5) |
| **Store Neighborhood** |  |  |  |  |
| **Demographics** |  |  |  |  |
| Non-Hispanic White residents |  |  |  |  |
| Q1 (0-28%) | ref | ref | ref | ref |
| Q2 (28-60%) | **2.7 (1.5-4.7)** | **2.0 (1.2-3.5)** | **1.7 (1.1-2.6)** | 1.5 (0.9-2.5) |
| Q3 (60-74%) | **2.9 (1.7-5.0)** | **1.8 (1.0-3.0)** | 1.5 (0.9-2.3) | 1.3 (0.8-2.2) |
| Q4 (74% +) | **3.6 (2.1-6.1)** | **2.3 (1.3-4.0)** | **1.6 (1.0-2.5)** | 1.2 (0.7-2.1) |
| Households under poverty |  |  |  |  |
| Q1 (0-7%) | ref | ref | ref | ref |
| Q2 (7-17%) | 1.1 (0.8-1.5) | 1.1 (0.8-1.4) | 1 (0.7-1.3) | 0.8 (0.6-1.1) |
| Q3 (17-26%) | **0.6 (0.4-0.9)** | 0.8 (0.5-1.2) | **0.6 (0.4-0.9)** | 0.7 (0.5-1.2) |
| Q4 (26% +) | 0.9 (0.6-1.2) | 1.1 (0.8-1.6) | 0.7 (0.5-1.0) | 0.9 (0.5-1.4) |
| Within .5 mile of school |  |  |  |  |
| No | ref | ref | ref | ref |
| Yes | **0.7 (0.6-0.9)** | 1.0 (0.8-1.3) | 1.1 (0.8-1.5) | 1.0 (0.7-1.4) |

### Disposable E-Cigarettes

In unadjusted analyses, compared with chain convenience stores, tobacco stores/vape stores were significantly more likely to carry disposable e-cigarettes, but other store types like grocery stores and pharmacies were less likely to have these products. These products were also more prevalent in NY stores compared with NJ stores and in stores in census tracts with the 2^nd^ and 4^th^ quartile of White residents compared with the lowest quartile. Stores in neighborhoods with the 3^rd^ quartile of residents under poverty compared with the lowest quartile were less likely to have disposable e-cigarettes available. In adjusted analyses, only store type was a significant correlate of disposable e-cigarette availability.

## Discussion

The availability of newer tobacco products like nicotine pouches and disposable e-cigarettes was found in roughly half the stores in our sample in some cases exceeding the availability of more established tobacco products such as e-cigarette pods and smokeless tobacco products. Availability varies in different parts of the country and patterns of availability of these two product types are not the same. Though these products share some common features such as relatively high nicotine concentration, flavor availability, ‘tobacco-free’ claims, and relatively low price,^16-19^ retail marketing appears to differ. Nicotine pouches seem to be more common in chain convenience stores likely driven by their promotion by major tobacco companies. They are also focused in neighborhoods with more non-Hispanic White residents--perhaps capitalizing on marketing to those who already use smokeless tobacco as non-Hispanic White populations use smokeless tobacco at higher rates than any other racial/ethnic group.^20^ In contrast, disposable e-cigarettes appear to be more prevalent in tobacco/vape shops, but not in other non-specialty store types like groceries and pharmacies. Encouragingly in our sample, there does not appear to be targeting of either product by neighborhood poverty or in stores near schools. Additional retail surveillance in more locations and especially outside of predominantly urban centers is needed to confirm such patterns.

## Conclusion

Our results provide an early indication of different patterns of retail availability of newer non-combustible tobacco products of nicotine pouches and disposable e-cigarettes. As both product types potentially face increased regulation under the FDA pre-market authorization process, understanding changes in their availability in the retail environment is critical to inform potential policies regulating their sale and marketing.

## Data Availability

All data produced in the present study are available upon reasonable request to the authors.

## Data Availability

All data produced in the present study are available upon reasonable request to the authors.

